# Warmer night-time temperatures are linked to poorer associative memory performance

**DOI:** 10.64898/2026.06.18.26356006

**Authors:** Anna Behler, Renate Thienel, Nicholas Bayliss, Felicity Simpson, Kerrie McAloney, Jessica Adsett, Nicholas G. Martin, Michael Breakspear, Michelle K. Lupton

**Affiliations:** School of Science, College of Engineering, Science and Environment, University of Newcastle, Australia; Hunter Medical Research Institute, Newcastle, Australia; School of Medicine and Public Health, College of Health, Medicine & Wellbeing, University of Newcastle, Australia; QIMR Berghofer Medical Research Institute, Brisbane, Australia; School of Biomedical Sciences, Faculty of Medicine, University of Queensland, Brisbane, Queensland, Australia; School of Biomedical Sciences, Faculty of Health, Queensland University of Technology, Brisbane, Queensland, Australia

**Author notes:** Corresponding Author: Dr Anna Behler, Hunter Medical Research Institute, Lot 1 Kookaburra Cct, New Lambton Heights NSW 2305, Australia.

## Abstract

Ambient temperature is emerging as an environmental factor that may influence cognitive performance in ageing populations. This is particularly relevant in Australia, where people live across diverse climatic regions spanning alpine to tropical conditions.

We examined daily temperatures and cognitive performance in 1,873 midlife and older adults (1,297 women, mean age 61.0 years) who completed the Creyos online battery (formerly Cambridge Brain Sciences). Twelve tasks assessed memory, visuospatial processing, language, attention, and executive function. Task scores were linked to postcode-level contemporaneous weather data. The scores were analysed in relation to maximum and minimum air and wet-bulb temperatures and postcode- and month-relative temperature percentiles. Regression models adjusted for age, sex, education, socioeconomic status, and climate zone, with season included for air and wet-bulb measures.

Higher minimum, but not maximum, temperature was associated with poorer performance on Paired Associates, a task assessing associative memory. This pattern was observed for air temperature, wet-bulb temperature, and temperature percentile, suggesting poorer memory performance after warmer nights, both in absolute terms and relative to local seasonal norms. Temperature was not significantly associated with performance on any other task, including measures of short-term/working memory, visuospatial processing, language, attention, or executive function.

These findings suggest a task-specific association between higher overnight temperature and poorer associative memory performance, rather than a general reduction in cognition. Further studies incorporating personal exposure and sleep measures are needed to clarify whether night-time thermal conditions affect cognitive health in midlife and older populations.

## INTRODUCTION

Ambient temperature has emerged as an important environmental exposure for cognitive performance (Yeganeh et al., 2018), particularly among older adults who may be more vulnerable to thermal stress (Byun et al., 2024). While the systemic physiological and health consequences of heat exposure are well established (Cramer et al., 2022; Ebi et al., 2021), less is known about its implications for brain health and cognitive function. This is an important gap because thermal stress may influence cognitive performance through physiological pathways relevant to brain function, including effects on metabolism, inflammatory processes and cardiovascular strain (Behler et al., 2026). Environmental heat may therefore affect cognition not only through general systemic strain, but also through pathways that are relevant to neural function. Understanding these links is increasingly important as populations experience more frequent and severe heat events (IPCC, 2023).

A growing body of population-based research suggests that environmental temperature is associated with cognitive performance, particularly in older adults. Studies from China, the United States, Taiwan, and Germany have linked higher ambient or residential temperatures with poorer global cognitive performance and domain-specific deficits (Dai et al., 2016; Hou & Xu, 2023; Lo et al., 2021; Yin et al., 2024; Zhao et al., 2021). For example, Yin et al. (2024) reported that extremely high temperatures were associated with reduced cognitive function across acute and longer-term exposure windows in China, while Hou and Xu (2023) found that higher monthly temperatures were associated with lower global cognitive scores in older Chinese adults. Studies in other regional settings suggest that cognitive performance may be affected by both heat and cold, although the direction and strength differ across populations (Dai et al., 2016; Khan et al., 2021; Zhao et al., 2021). These regional differences are important because the generalisability of existing findings remains uncertain and the same temperatures may not have equivalent cognitive implications across climatic contexts. Existing studies also differ in the cognitive batteries used and in whether they assess global cognition or domain-specific performance. Zhao et al. (2021), in a women-only sample using the Consortium to Establish a Registry for Alzheimer’s Disease (CERAD) neuropsychological assessment battery, reported the clearest association for episodic memory, with weaker or less consistent associations for semantic memory, constructional praxis, and executive function. In contrast, Hou and Xu (2023), using the Mini Mental State Examination (MMSE) subdomains in a mixed-gender sample, reported associations across several domains, including general ability, reaction ability, attention and calculation, memory, language comprehension and self-coordination.

A further unresolved issue is the functional nature of the temperature–cognition association. Several studies suggest that the relationship may not be strictly linear. For example, Dai et al. (2016) reported higher odds of low global cognitive performance at both lower and higher residential temperatures, while Zhao et al. (2021) observed an inverse U-shaped association between ambient temperature and cognitive function in elderly women, with the highest cognitive scores at approximately 15°C/59°F. Some research has also suggested that cumulative or repeated heat exposure may further compound risk, with heatwave exposure associated with increased risk of cognitive impairment and faster cognitive decline in some older populations (Choi et al., 2023; Ye et al., 2024; Zhou et al., 2023). Together, these findings suggest that both heat and cold exposure may be relevant for cognition, and that linear models alone may not fully capture temperature-related cognitive effects.

The generalisability of existing findings to the Australian population remains uncertain. Population-based studies of temperature and cognition have largely been conducted in North America, Europe, and Asia, where climatic conditions, housing characteristics, infrastructure, population acclimatisation, and behavioural responses to heat differ from the Australian context. Australia provides a particularly relevant setting because of its substantial climatic heterogeneity. Coastal Australia spans tropical, subtropical, temperate, and cooler climatic zones, while inland areas include extensive arid and semi-arid environments. As a result, the same absolute temperature may have different cognitive implications depending on local climate, built environment, and prior acclimatisation (Wijayanto et al., 2017). Australia has also warmed substantially since national weather records began, with increases in both the frequency and intensity of extreme heat events (Bureau of Meteorology & CSIRO, 2024).

The present study addresses these gaps by examining associations between environmental temperature and cognitive performance in a large Australian adult cohort. We linked online cognitive assessment data across 12 cognitive tasks with postcode-level daily weather exposure derived from a weather database. We tested whether cognitive performance was associated with absolute daily maximum and minimum air temperature, wet-bulb temperature (that accounts for both heat and humidity), and region- and month-specific temperature percentiles. Given previous evidence for non-linear temperature–cognition associations, we evaluated both linear and quadratic exposure–response models. We hypothesised that higher daily temperatures would be associated with poorer cognitive performance, and that associations may differ according to exposure definition and cognitive domain.

## METHODS

### Participants

Participants were drawn from the Prospective Imaging Study of Ageing: Genes, Brain and Behaviour (PISA) (Lupton et al., 2021). PISA is a large cohort study of midlife and older adults. For the PISA online component, surveys and cognitive testing were used to characterise an Australia-wide sample. PISA participants were recruited from those who were previously enrolled in genetic epidemiology studies at QIMR Berghofer (detailed in Lupton et al., 2022). The recruitment pool comprised participants with existing genome-wide genotype data from QIMR Berghofer cohorts, including Australian twin and family studies and genome-wide association studies of physical and psychiatric traits. There were no exclusions based on any health condition.

The comprehensive PISA online survey assessed key demographics, education, occupation and medical history among other topics, as detailed in Lupton et al. (2021) and included online cognitive testing (Thienel et al., 2024). Demographics included date of birth, education level and residential postcode. PISA online participants were included in this study if they had completed at least one of the cognitive tests and had supplied their age, years of education and a valid residential postcode, resulting in a total of 2,247 participants. Years of education were derived from highest level of tertiary education. The socio-economic index (SEI) was derived from the participant’s occupation using the Australian Socioeconomic Index 2006 (AUSEI06) scale (McMillan et al., 2009). Participant sex was derived from genome-wide genotype data as part of standard genetic quality-control procedures. The participants took part between the 3^rd^ of March 2018 and the 20^th^ of January 2022.

Ethical approval was obtained from the Human Research Ethics committee of QIMR Berghofer (P2210) and the Human Research Ethics committee of the University of Newcastle (H-2020-0439).

### Cognitive Measures

Cognition was assessed using the Creyos online battery (https://creyos.com/), previously known as Cambridge Brain Sciences. The tasks were undertaken unsupervised in the participant’s home environment. The battery consists of twelve self-administered tasks that evaluate memory, visuospatial, language and executive function (Corbett et al., 2015). The full battery takes approximately 30 minutes to complete.

Episodic memory was assessed with the Paired Associates test, where participants learn and recall associations between objects and their locations. The Rotations Test assesses mental rotation and visuospatial processing by asking participants to decide if two rotated 3D shapes are identical. Polygons measure visuospatial processing where the user judges whether overlapping shapes match a target polygon. Feature Match measures focused attention by asking participants to decide if two complex images have matching features. Grammatical Reasoning requires verbal reasoning by verifying statements about the spatial relationships between simple shapes and Digit Span is based on the Wechsler Adult Intelligence Scale-Revised (Wechsler, 1981), but here only includes the forward condition. Therefore, it only assesses attention and short-term memory. Token Search is based on a test used to measure strategy-based problem-solving during search behaviour. Double Trouble is a variant of the Stroop test (Stroop, 1935) that assesses response inhibition and selective attention, and Odd One Out measures deductive reasoning by requiring users to identify the rule that differentiates one shape or pattern from the others in a group. Monkey Ladder requires clicking boxes in ascending numerical order testing visuospatial working memory, Spatial Span tests spatial attention/short-term memory by repeating a sequence of flashing squares (based on the Corsi block tapping task) and Spatial Planning assesses planning and executive function, heavily based on the classic “Tower of London” puzzle.

### Historical weather data

Daily weather data were obtained from the SILO (Scientific Information for Land Owners) climate database, an Australian gridded climate archive based on Bureau of Meteorology observations (Jeffrey et al., 2001). SILO provides spatially and temporally complete daily climate surfaces for Australia from 1889 onwards on a 0.05° × 0.05° grid (approximately 5 × 5 km). Gridded datasets are generated by spatial interpolation of observational weather station data. SILO includes 19 climate variables overall: 18 daily variables and monthly rainfall. In the present analysis, we used four SILO variables: daily maximum air temperature, daily minimum air temperature, relative humidity at the time of maximum air temperature, and relative humidity at the time of minimum air temperature. Daily minimum and maximum temperatures follow the Bureau of Meteorology 9am observation convention, which represents the minimum and maximum temperatures observed over the relevant preceding 24-hour observation periods. The daily maximum temperature is the highest temperature in the 24 hours before the 9am observation and is assigned to the previous calendar day. The daily minimum temperature is the lowest temperature in the 24 hours before the 9am observation and is assigned to the day of observation. The relative humidity variables correspond to humidity at the time of daily maximum and minimum temperature, respectively. We further derived monthly percentile ranks for maximum and minimum temperature relative to a postcode-specific 1990–2020 historical baseline.

### Mapping weather data and postcode areas

We mapped participant postcodes to gridded SILO data using the Australian Bureau of Statistics (ABS) Postal Areas (POA) digital boundary files from the Australian Statistical Geography Standard (ASGS) Edition 3, Non-ABS Structures, 2021 release. Postal Areas are ABS-defined approximations of Australia Post postcodes and are provided as polygon shapefiles representing the spatial extent postcode regions across Australia. These POA boundaries are designed for statistical and spatial analysis. In this study, we used the 2021 POA shapefile as the national postcode boundary layer, retained all valid numeric postcode areas, and reprojected the geometries to align with the SILO climate grid. The resulting polygon layer formed the spatial basis for assigning postcode areas to SILO grid cells.

All numeric POAs with valid geometry were rasterized onto the SILO grid using an area-based assignment, where each postcode polygon was overlaid on the SILO grid and every grid cell touched by the polygon boundary or interior was assigned to that postcode, rather than representing the postcode by a single point location. This preserves the spatial footprint of larger postcode regions and allows downstream climate estimates to reflect the full set of SILO cells intersecting the postcode area. POAs occupying more than one SILO grid cell were retained in a gridded mask, while POAs occupying zero or one raster cell were instead represented by a centroid-based nearest-grid point.

### Mapping weather to cognitive assessment

Weather exposure was assigned to each cognitive assessment record using the participant’s residential postcode and the date of test completion. For each assessment date, postcode-level values were extracted for daily maximum air temperature *T*_max_, daily minimum air temperature *T*_min_, relative humidity at the time of maximum temperature, and relative humidity at the time of minimum temperature. We refer to maximum and minimum air temperature variables as dry-bulb temperature, that is, the conventional ambient air temperature measured independently of humidity. Wet-bulb temperatures at the time of daily maximum and minimum air temperature (*T*_w,max_ and *T*_w,min_) were then calculated using the Stull approximation (Stull, 2011) to capture heat exposure after accounting for evaporative cooling potential.

In addition to absolute weather conditions, we derived month-specific percentile ranks for daily maximum and minimum temperature (*T*_max,percentile_ and *T*_min,percentile_) relative to a postcode-specific historical baseline spanning 1990 to 2020. A binary heatwave indicator was also generated, defined as three consecutive days comprising the assessment day and the two preceding days during which mean daily temperature exceeded both the postcode- and month-specific 90th percentile of the historical baseline and an absolute threshold of 27 °C.

### Season and climate zone

Residential postcodes were also classified into climate zones defined by the National Construction Code (NCC) to adjust for broad regional climate differences. The NCC classification distinguishes eight broad climatic regions: zone 1, characterised by hot humid summers and warm winters; zone 2, with warm humid summers and mild winters; zone 3, with hot dry summers and warm winters; zone 4, with hot dry summers and cool winters; zone 5, representing warm temperate regions; zone 6, representing mild temperate regions; zone 7, representing cool temperate regions; and zone 8, representing alpine regions. Climate-zone assignments were obtained primarily from the Australian Energy Regulator postcode climate-zone lookup (Australian Energy Regulator, n.d.). Where postcode assignments were missing or ambiguous, we used postcode-level information from the Nationwide House Energy Rating Scheme as a supplementary source to identify the most appropriate corresponding NCC climate zone (Nationwide House Energy Rating Scheme, n.d.).

Assessment season was derived from the cognitive testing date using Southern Hemisphere seasons: summer, December-February; autumn, March-May; winter, June-August; and spring, September-November.

### Statistical analysis of linear temperature associations

Associations between ambient temperature and cognitive performance were estimated using ordinary least squares regression. Separate models were fitted for each combination of 12 z-standardised cognitive outcomes and six temperature exposures, giving 72 primary exposure-outcome models. The temperature exposures were daily maximum temperature *T*_max_, daily minimum temperature *T*_min_, wet-bulb temperature at the time of daily maximum temperature *T*_w,max_, wet-bulb temperature at the time of daily minimum temperature *T*_w,min_, maximum-temperature percentile *T*_max,percentile_, and minimum-temperature percentile *T*_min,percentile_. For absolute temperature and wet-bulb models, the primary model was,

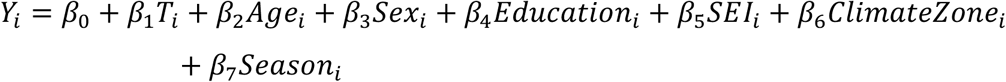

For percentile-based models, the same specification was used,

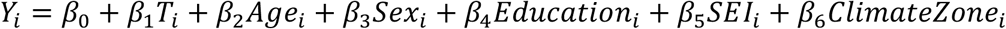

Covariates were selected *a priori* to account for demographic, socioeconomic, and regional factors that may be associated with both cognitive performance and environmental temperature exposure. All models adjusted for age, sex, years of education, SEI, and climate zone. SEI was included to account for socioeconomic differences that may influence both cognitive performance and residential location. Climate zone was modelled as a categorical covariate (with zone 7 as the reference category) to adjust for broad regional climatic differences across Australia. For absolute dry-bulb and wet-bulb temperature exposures, assessment season (Winter as reference season) was additionally included to account for seasonal variation. However, season was not included in percentile-based models because these exposure variables were already normalised within postcode and calendar month.

Regression coefficients for temperature exposures represent the adjusted difference in standardised cognitive performance associated with a 1 °C increase in absolute or wet-bulb temperature, or with a one-percentile-point increase in percentile-based temperature exposure. Heteroscedasticity-consistent standard errors were used for inference using the HC3 correction (MacKinnon & White, 1985). To account for multiple testing across the 72 primary exposure–outcome models, p-values were adjusted using the Benjamini–Hochberg False Discovery Rate (FDR) procedure (Benjamini & Hochberg, 1995).

### Assessment of non-linear temperature effects

To test whether temperature-cognition associations were better captured by non-linear effects, linear and quadratic specifications were compared for each cognitive outcome and temperature exposure. For the quadratic models, the exposure variable was mean-centred before squaring to reduce collinearity between the linear and quadratic terms. Quadratic models used the same covariate structure as the corresponding linear models, including season for absolute (dry- and wet-bulb) temperature exposures but not for percentile exposures:

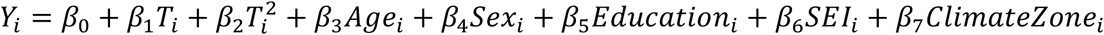

Model comparison between the linear and quadratic model was based on Akaike’s Information Criterion (AIC) from the conventional non-robust ordinary least squares fits, because AIC is likelihood-based and already penalises the inclusion of additional parameters. The quadratic specification was considered to provide meaningful improvement only when its AIC was more than two units lower than that of the corresponding linear model; otherwise, the linear specification was retained.

### Latent factors of cognition

Consistent with dimensionality-reduction approaches previously applied to PISA online cognitive data (Faucher et al., 2024; Thienel et al., 2024), we used exploratory factor analysis to derive broader cognitive factors from the 12 task-level outcomes. This shared variance among cognitive tasks as latent cognitive domains as an exploratory data-driven approach rather than a confirmatory test of a pre-specified cognitive factor structure. Only participants who completed all 12 cognitive tests were included for exploratory factor analysis. Suitability for factor analysis was assessed using Bartlett’s test of sphericity and the Kaiser-Meyer-Olkin measure of sampling adequacy.

The number of factors was guided by the eigenvalue structure of the cognitive-task correlation matrix and scree-plot inspection. Factors were extracted using principal factor analysis with oblimin rotation, allowing the factors to correlate. Because factor orientations are arbitrary, each factor was oriented so that the task with the largest absolute loading had a positive loading. Factor scores were standardised to approximately mean zero and unit variance.

Factor scores were then analysed using the same linear temperature-regression framework as the primary task-level models. Separate models were fitted for each retained factor and each of the six temperature exposures. Models adjusted for age, sex, years of education, SEI, and climate zone, with season included only for absolute dry-bulb and wet-bulb exposures. HC3 robust standard errors were used for inference. FDR correction was applied globally across the 18 factor-level exposure tests.

## RESULTS

### Participant and temperature exposure characteristics

Of the 2,247 eligible participants, 374 were excluded because temperature exposure could not be assigned, leaving a final sample of 1,873. Of these, 1,297 (69.2%) identified as female and 576 (30.8%) as male. Mean age was 61.0 ± 6.7 years (range: 42–75 years). The mean total years of education was 13.4 ± 2.9 years (range: 0–25). The mean SEI was 62.9 ± 23.7 (range: 1–100). Sample sizes varied across cognitive tests because not all participants completed every task. Test-specific sample sizes ranged from 1,492 for Paired Associates to 1,768 for Odd One Out. Full sample sizes for each task are provided in Supplementary Table S2. These participants lived in 765 unique postcode areas spanning all Australian states and territories (Fig. 1A).

**Figure 1.**
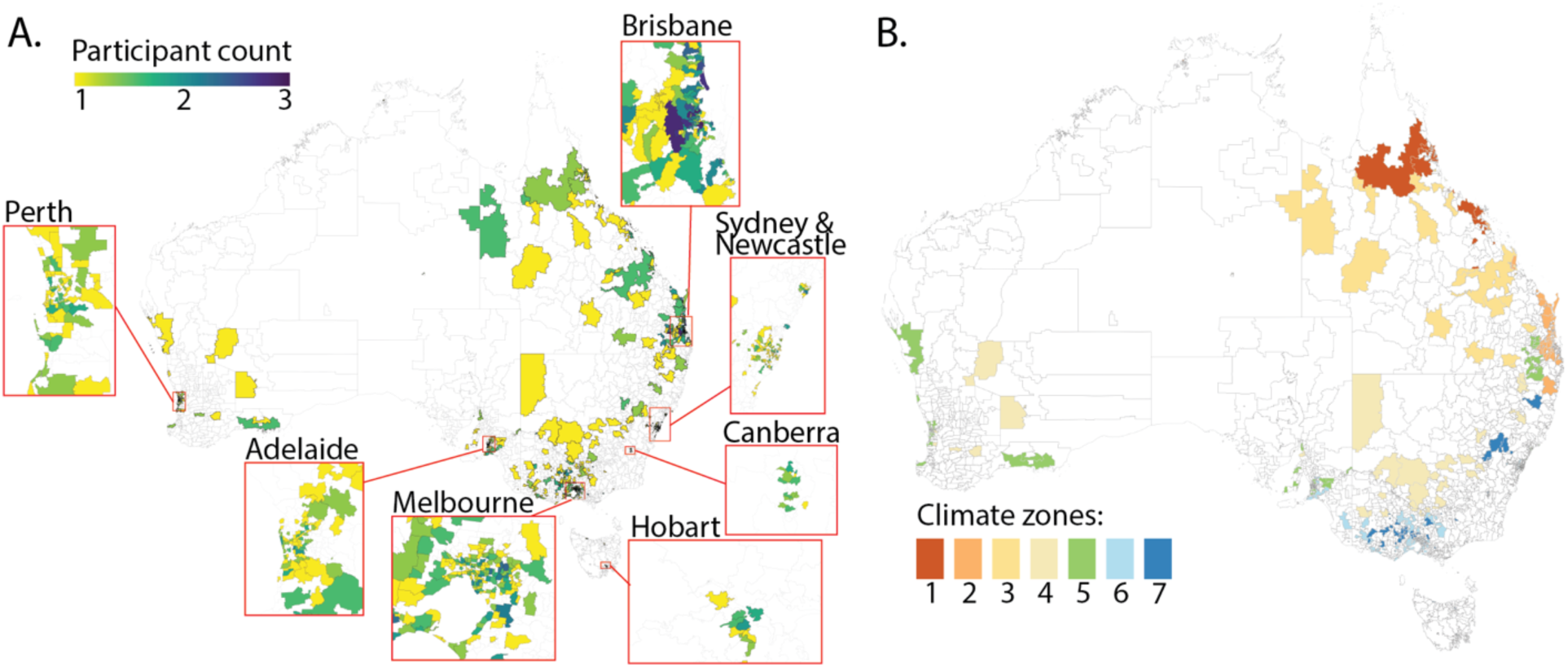
A. Distribution of participants across Australia. B. Climate zones of postcode areas included.

Participants completed the tasks across all four seasons, with a broadly even distribution: spring (*n* = 600, 32.0%), summer (*n* = 457, 24.4%), winter (*n* = 422, 22.5%), and autumn (*n* = 394, 21.0%). Participants were distributed across seven of the eight Australian climate zones (Fig. 1B), with the majority residing in zone 2 (warm humid summer/mild winter; 35.9%) and zone 6 (mild temperate; 26.6%), followed by zone 5 (warm temperate; 9.0%) and zone 7 (cool temperate; 8.5%). Zones 1 (hot humid summer, warm winter), 3 (Hot dry summer, warm winter), and 4 (Hot dry summer, cool winter) each accounted for fewer than 3% of the sample. Detailed climate-zone-by-season sample sizes are provided in Supplementary Table S3.

Figure 2 shows the distribution of temperature variables on the day of testing. Daily maximum temperatures *T*_max_ ranged from 8.5 to 44.6°C (*M* = 25.3°C, *SD* = 6.5°C) and daily minimum temperatures *T*_min_ from –4.5°C to 29.0°C (*M* = 14.2°C, *SD* = 5.9°C). Temperature percentiles were *T*_max,percentile_ = 51.4 ± 31.0 and *T*_min,percentile_ = 50.1 ± 31.6. Wet-bulb temperatures were lower than dry-bulb equivalents, as expected, with *T*_w,max_ ranging from 4.72°C to 27.73°C (*M* = 17.65°C, *SD* = 4.79°C) and *T*_w,min_ from -4.66°C to 25.76°C (*M* = 12.93°C, *SD* = 5.72°C). Only 18 observations (1.0%) coincided with a heatwave day, precluding separate analysis of heatwave effects. Temperature distributions across seasons and climate zones are shown in Supplementary Figure S1.

**Figure 2.**
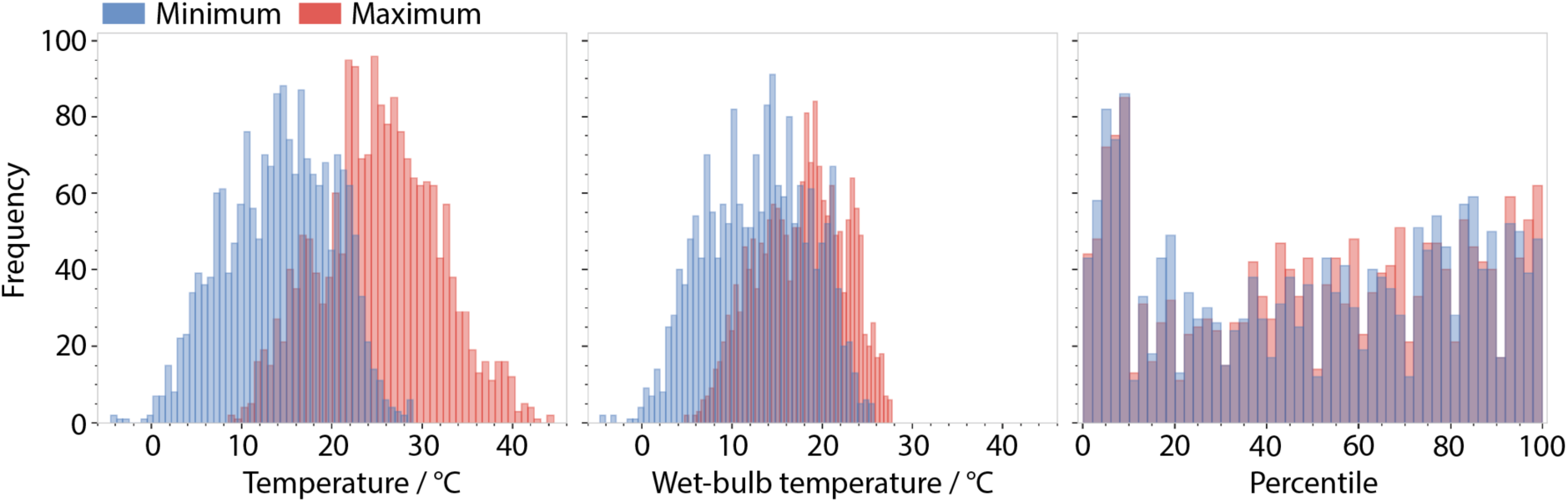
Distribution of environmental temperature at date of assessment.

### Linear Versus Quadratic Model Comparison

We first compared linear and quadratic temperature models for each cognitive outcome and exposure metric. The linear specification was retained for the primary analyses because quadratic models showed little evidence of improved fit. Across the six temperature metrics, the quadratic model was preferred by AIC in only one of 72 comparisons: Odd One Out for *T*_min,percentile_. However, this isolated quadratic term was not significant after FDR correction (Supplementary Table S4).

### Temperature-cognitive associations

Figure 3 shows the estimated associations between each temperature variable and each cognitive test. Coefficients represent the change in standardised cognitive performance per unit increase in the exposure, adjusted for age, sex, years of education, socio-economic index, climate zone, and, for absolute temperature variables, assessment season. Across the 72 models, three associations, all involving the Paired Associates test, survived FDR corrections. The remaining eleven tests were not significantly associated with any temperature variable (Supplementary Table S5).

**Figure 3.**
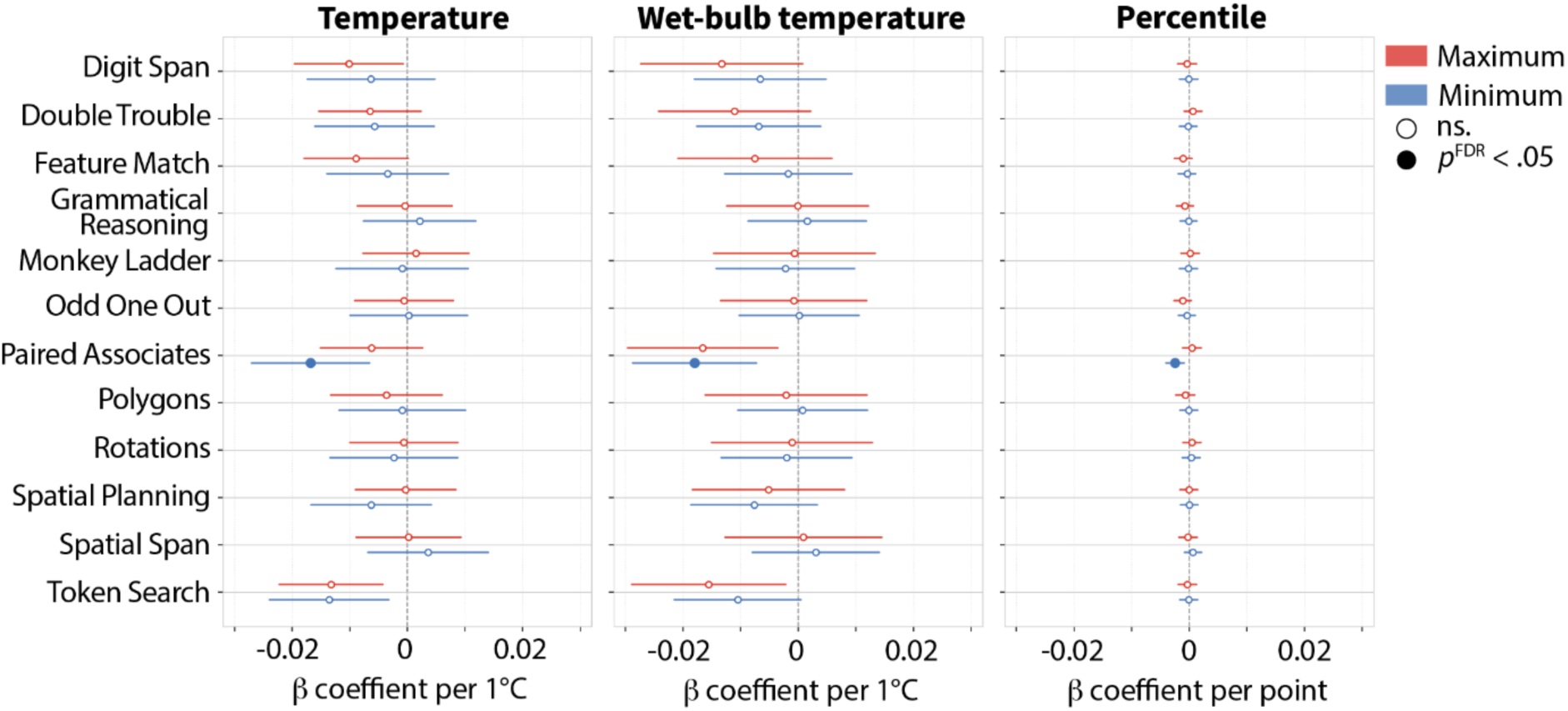
Forest plot showing β coefficients and 95% confidence intervals for associations between temperature exposures and standardised cognitive test performance. Filled markers indicate associations surviving global False Discovery Rate (FDR) correction; open markers indicate non-significant (n.s.) associations.

For Paired Associates, all three significant associations reflected poorer performance with higher minimum-temperature exposure (Fig. 4). Lower Paired Associates performance was associated with higher *T*_min_ (*b* = –0.017, 95% CI [–0.027, – 0.007], *p* = .001, *p*^FDR^ = .046), as well as *T*_w,min_ (*b* = –0.018, 95% CI [–0.029, –0.007], *p* = .001, *p*^FDR^ = .046) and *T*_min,percentile_ (*b* = –0.002 per percentile point, 95% CI [–0.004, –0.001], *p* = .002, *p*^FDR^ = .048).

**Figure 4.**
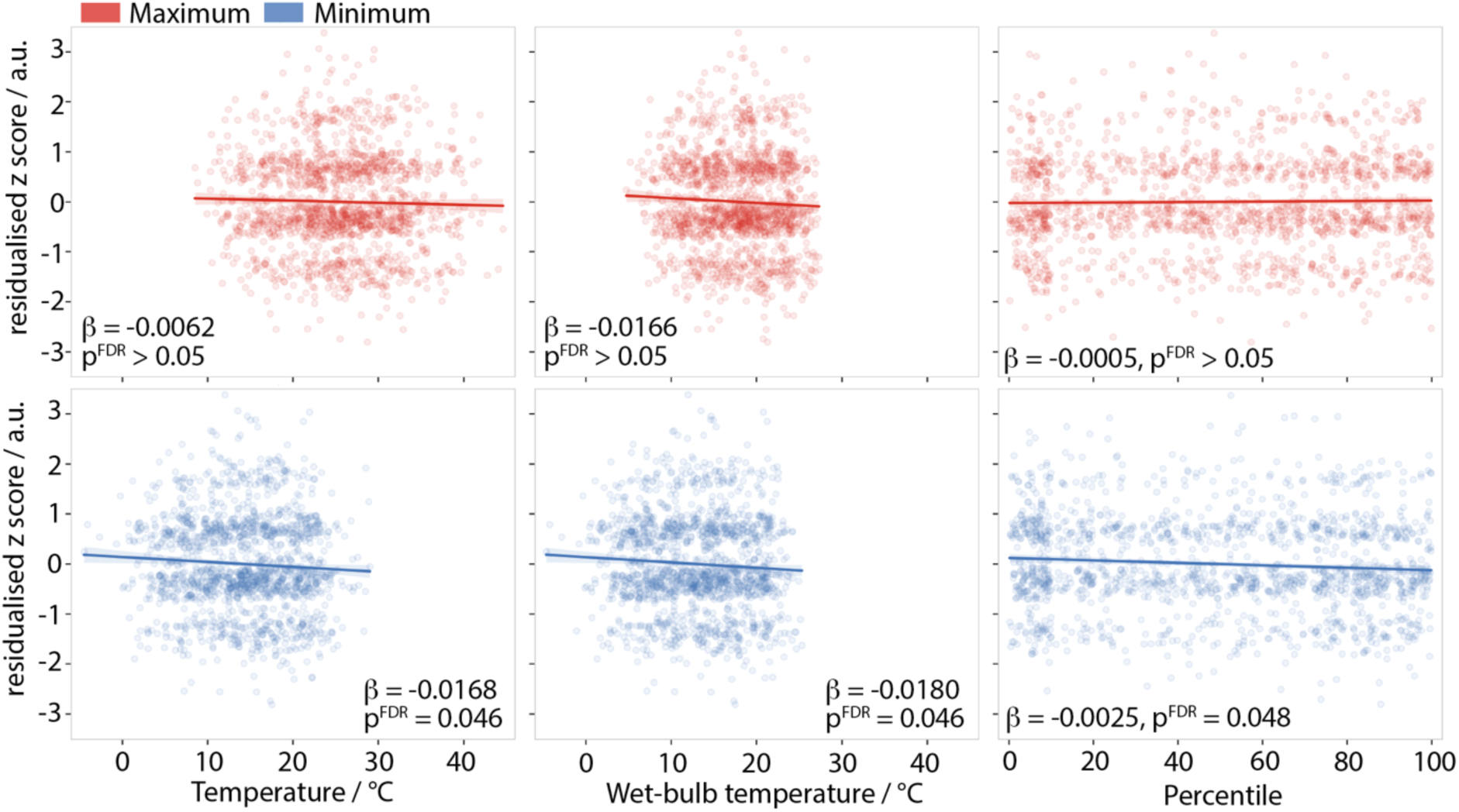
Paired Associates performance across minimum-temperature exposures. Adjusted Paired Associates z-scores as a function of daily minimum temperature, wet-bulb minimum temperature, and minimum temperature percentile. Shaded bands indicate 95% confidence intervals.

### Demographic, socioeconomic, seasonal and regional covariates

Age, years of education, socioeconomic index, and sex were included as adjustment variables in all models; their associations with cognitive performance were broadly consistent with expected demographic patterns. Seasonal and regional covariate associations were present but task-specific. Across absolute-temperature models, Digit Span scores were higher in summer than winter (*b* = 0.218–0.261, *p* = .004–.015), and Token Search scores were higher in both summer (*b* = 0.204–0.244, *p* = .005–.024) and autumn (*b* = 0.227–0.253, *p* = .002–.005). Double Trouble scores were lower in spring relative to winter (*b* = –0.185 to –0.172, *p* = .009–.018). Paired Associates performance was lower in climate zone 6 (mild temperate) than zone 7 (cool temperate) across all exposure specifications (*b* = –0.240 to –0.192, *p* = .013–.048), with a similar but less consistent pattern for zone 5 (warm temperate; *b* = –0.230 to –0.170, *p* = .019–.084). Token Search performance was higher in climate zone 4 (hot dry summer, cool winter) than zone 7 in absolute-temperature models (*b* = 0.304–0.376, *p* = .013–.038), but not in temperature percentile-based models.

### Temperature effects across latent cognitive factors

We extracted three cognitive factors from the standardised task scores (for full details, see Supplementary Material). The factor structure was broadly interpretable as Span/Memory, Visuospatial/Reasoning, and Verbal/Executive. Temperature exposure was not significantly associated with any of these three factor scores (Supplementary Table S1). These findings suggest that the task-level Paired Associates association was not explained by a broader factor-level association across the cognitive battery.

## DISCUSSION

In our sample of midlife and older Australians, higher minimum-temperature on the day of assessment showed a highly specific association with poorer performance on Paired Associates, a task indexing associative and episodic memory. This association was observed for minimum air temperature, minimum wet-bulb temperature, and minimum-temperature percentile, suggesting an association both in absolute terms and relative to local seasonal norms. By contrast, no other cognitive task survived correction for multiple comparisons, temperature was not associated with broader latent cognitive factors, and quadratic models did not meaningfully improve model fit. Overall, the findings suggest a selective association between higher daily minimum temperatures and performance on an associative memory task, rather than a general reduction in cognitive performance across the battery.

The Creyos Paired Associates task is distinct in that it specifically assesses episodic memory, requiring individuals to form and recall associations between an item (an icon) and its spatial location (a hidden box). Unlike other Creyos tasks that predominantly assess short-term memory, reasoning, attention, or visual processing abilities, the Paired Associates task relies on the encoding and retrieval of specific item–location (“what-and-where”) associations, a core feature of episodic memory functioning. Impairment in episodic memory is the hallmark cognitive symptom of Alzheimer’s Disease (AD) - often one of the earliest and most clinically significant manifestations of the condition. Consequently, episodic memory outcomes are frequently used as primary endpoints in the evaluation of disease-modifying, symptomatic, and risk-reduction interventions targeting AD (Dubois et al., 2007).

Our findings refine previous population-based evidence linking ambient temperature with cognition. Several studies have reported associations between non-optimal temperature and poorer global cognition, cognitive impairment, or domain-specific cognitive deficits (Dai et al., 2016; Hou & Xu, 2023; Khan et al., 2021; Lo et al., 2021; Yin et al., 2024; Zhao et al., 2021). Our findings are consistent with this broader literature in showing an adverse association between warmer environmental conditions and cognition, but they suggest a narrower effect in this Australian cohort. Importantly, the domain-specific pattern aligns with Zhao et al. (2021), who reported the clearest temperature-related association for episodic memory in an older women-only cohort, with weaker or less consistent associations across other cognitive domains. In the present study, the association was similarly confined to a task requiring participants to learn and recall object–location pairings. Together, these findings raise the possibility that episodic or associative memory may be a particularly sensitive cognitive endpoint for short-term temperature exposure, although replication in cohorts with repeated cognitive testing and personal exposure measurement is needed before strong domain-specific conclusions can be drawn.

The finding that minimum-temperature exposure, rather than maximum temperature, was most consistently associated with memory performance may point to the importance of night-time thermal conditions. Warmer nights can reduce opportunities for physiological recovery and may disrupt sleep. This interpretation is supported by evidence that higher night-time temperatures are associated with shorter or poorer sleep (Bigler & Janzen, 2024; Minor et al., 2022; Obradovich et al., 2017), and by evidence that sleep restriction impairs memory formation (Crowley et al., 2024). Because the Paired Associates task depends on learning and recall, it is plausible that this task would be sensitive to sleep-related effects of warmer nights. However, sleep quality immediately prior to the cognitive test completion was not measured in the present study, so this mechanism remains speculative.

The finding is relevant in the context of climate change because very high night-time temperatures have become more frequent in Australia and are recognised as an important contributor to heat stress (Bureau of Meteorology & CSIRO, 2024). Cooler nights allow recovery from daytime heat, whereas high overnight temperatures prolong heat exposure and reduce physiological recovery. The present finding that Paired Associates performance was most consistently associated with minimum-temperature exposure therefore suggests that warmer nights may be relevant to memory performance in midlife and older populations. This may be particularly important for older adults, who are more vulnerable to heat because of age-related changes in thermoregulation, sweating, skin blood flow, hydration, chronic disease burden, and medication use (Meade et al., 2020; Millyard et al., 2020; Wee et al., 2023). However, heat-related cognitive vulnerability is not determined by temperature alone. Vulnerability also appears to be socially variable, as shown in a study conducted in the United States, where cumulative extreme heat exposure was associated with faster cognitive decline. This effect was particularly evident among Black older adults and residents of socioeconomically disadvantaged neighbourhoods (Choi et al., 2023). Evidence from older Chinese adults suggests that heatwave-related cognitive risk is modified by air pollution and green space (Zhou et al., 2023), indicating that cognitive outcomes may be better protected where environmental and adaptive factors reduce effective heat exposure.

The convergence between dry-bulb minimum temperature, wet-bulb minimum temperature, and minimum-temperature percentile strengthens the interpretation that the result was not driven by a single exposure definition. Wet-bulb temperature combines air temperature and humidity, and is widely used as an indicator of humid heat stress because it reflects limits on evaporative cooling more directly than temperature alone (Raymond et al., 2020; Sherwood & Huber, 2010). By contrast, percentile-based temperature metrics capture whether a given temperature is unusually warm relative to the local climate. Similar approaches are commonly used in heatwave and heat-health research to account for regional climatic adaptation and local temperature distributions (Gasparrini et al., 2015; Nairn & Fawcett, 2014).The finding across these related but distinct measures suggests that both absolute thermal load and locally unusual minimum temperatures may be relevant to memory performance.

There was no evidence for non-linear temperature–cognition associations. This contrasts with studies in the New England region in North America and Germany, which reported associations between temperature and cognition (Dai et al., 2016; Zhao et al., 2021). The present Australian sample had a limited range of lower temperatures, reducing our ability to detect cold-related cognitive decrements. Although some assessment days included low minimum temperatures, the exposure distribution was centred on mild-to-warm conditions, and most participants lived in warm humid, warm temperate, or mild temperate climate zones. Therefore, the absence of quadratic effects should not be interpreted as evidence that cold exposure is irrelevant for cognition, but rather that linear models adequately described the temperature range captured in this cohort.

Several limitations should be considered. Temperature exposure was assigned using postcode-level outdoor weather data, rather than personal or indoor temperature measurements. We therefore do not know whether participants completed the online cognitive tasks in air-conditioned environments, naturally ventilated homes, workplaces, or other indoor settings. This is important in Australia, where air-conditioning penetration is high and cooling is a common heat-adaptation strategy (Zander et al., 2021), although access and use are not universal and may depend on housing quality, affordability, energy costs, and individual behaviour. In addition, some Australian postcode areas are geographically large, which may introduce exposure misclassification. The sample also included an over-representation of women, which may limit generalisability and reduce precision for estimating associations in men. Although models were adjusted for sex, the uneven sex distribution may have limited our ability to detect sex-specific temperature effects or interactions.

In conclusion, higher overnight temperatures were associated with poorer episodic memory performance in middle-aged and older Australians, while other cognitive domains showed no robust associations. Our findings have potential public-health significance because episodic memory is central to cognitive ageing, and because minimum-temperature exposure may become increasingly relevant as warmer nights reduce opportunities for physiological recovery. Future studies that combine repeated cognitive assessments with personal or indoor exposure monitoring and sleep measurements would be important for clarifying the mechanisms linking warmer thermal conditions to cognitive performance.

## Data Availability

All data produced in the present study are available upon reasonable request to the authors

## References

1. Australian Energy Regulator. (n.d.). *ACIL Allen Summary of benchmarks 2017—Updated 27 March* 2018. Retrieved June 14, 2025, from https://www.aer.gov.au/documents/acil-allen-summary-benchmarks-2017-updated-27-march-2018

2. Behler, C., Fay, M., Ramadan, S., Breakspear, M., & Behler, A. (2026). Neurophysiology of brain temperature dysregulation in humans. Journal of Neurophysiology, jn.00418.2025. 10.1152/jn.00418.2025

3. Benjamini, Y., & Hochberg, Y. (1995). Controlling the False Discovery Rate: A Practical and Powerful Approach to Multiple Testing. Journal of the Royal Statistical Society Series B: Statistical Methodology, 57(1), 289–300. 10.1111/j.2517-6161.1995.tb02031.x

4. Bigler, P., & Janzen, B. (2024). Too hot to sleep. Journal of Environmental Economics and Management, 128, 103063. 10.1016/j.jeem.2024.103063

5. Bureau of Meteorology, & CSIRO. (2024). State of the Climate 2024. CSIRO and Bureau of Meteorology.

6. Byun, G., Choi, Y., Foo, D., Stewart, R., Song, Y., Son, J.-Y., Heo, S., Ning, X., Clark, C., Kim, H., Michelle Choi, H., Kim, S., Kim, S.-Y., Burrows, K., Lee, J.-T., Deziel, N. C., & Bell, M. L. (2024). Effects of ambient temperature on mental and neurological conditions in older adults: A systematic review and meta-analysis. Environment International, 194, 109166. 10.1016/j.envint.2024.109166

7. Choi, E. Y., Lee, H., & Chang, V. W. (2023). Cumulative exposure to extreme heat and trajectories of cognitive decline among older adults in the USA. Journal of Epidemiology and Community Health, 77(11), 728–735. 10.1136/jech-2023-220675

8. Corbett, A., Owen, A., Hampshire, A., Grahn, J., Stenton, R., Dajani, S., Burns, A., Howard, R., Williams, N., Williams, G., & Ballard, C. (2015). The Effect of an Online Cognitive Training Package in Healthy Older Adults: An Online Randomized Controlled Trial. Journal of the American Medical Directors Association, 16(11), 990–997. 10.1016/j.jamda.2015.06.014

9. Cramer, M. N., Gagnon, D., Laitano, O., & Crandall, C. G. (2022). Human temperature regulation under heat stress in health, disease, and injury. Physiological Reviews, 102(4), 1907–1989. 10.1152/physrev.00047.2021

10. Crowley, R., Alderman, E., Javadi, A.-H., & Tamminen, J. (2024). A systematic and meta-analytic review of the impact of sleep restriction on memory formation. Neuroscience & Biobehavioral Reviews, 167, 105929. 10.1016/j.neubiorev.2024.105929

11. Dai, L., Kloog, I., Coull, B. A., Sparrow, D., Spiro, A., Vokonas, P. S., & Schwartz, J. D. (2016). Cognitive function and short-term exposure to residential air temperature: A repeated measures study based on spatiotemporal estimates of temperature. Environmental Research, 150, 446–451. 10.1016/j.envres.2016.06.036

12. Dubois, B., Feldman, H. H., Jacova, C., DeKosky, S. T., Barberger-Gateau, P., Cummings, J., Delacourte, A., Galasko, D., Gauthier, S., Jicha, G., Meguro, K., O’Brien, J., Pasquier, F., Robert, P., Rossor, M., Salloway, S., Stern, Y., Visser, P. J., & Scheltens, P. (2007). Research criteria for the diagnosis of Alzheimer’s disease: Revising the NINCDS–ADRDA criteria. The Lancet Neurology, 6(8), 734–746. 10.1016/S1474-4422(07)70178-3

13. Ebi, K. L., Capon, A., Berry, P., Broderick, C., De Dear, R., Havenith, G., Honda, Y., Kovats, R. S., Ma, W., Malik, A., Morris, N. B., Nybo, L., Seneviratne, S. I., Vanos, J., & Jay, O. (2021). Hot weather and heat extremes: Health risks. The Lancet, 398(10301), 698–708. 10.1016/S0140-6736(21)01208-3

14. Faucher, C., Borne, L., Behler, A., Paton, B., Giorgio, J., Fripp, J., Thienel, R., Lupton, M. K., & Breakspear, M. (2024). A central role of sulcal width in the associations of sleep duration and depression with cognition in mid to late life. Sleep Advances, 5(1), zpae058. 10.1093/sleepadvances/zpae058

15. Gasparrini, A., Guo, Y., Hashizume, M., Lavigne, E., Zanobetti, A., Schwartz, J., Tobias, A., Tong, S., Rocklöv, J., Forsberg, B., Leone, M., De Sario, M., Bell, M. L., Guo, Y.-L. L., Wu, C., Kan, H., Yi, S.-M., De Sousa Zanotti Stagliorio Coelho, M., Saldiva, P. H. N., … Armstrong, B. (2015). Mortality risk attributable to high and low ambient temperature: A multicountry observational study. The Lancet, 386(9991), 369–375. 10.1016/S0140-6736(14)62114-0

16. Gomez, L. M., Mitchell, B. L., McAloney, K., Adsett, J., Garden, N., Wood, M., Diaz-Torres, S., Garcia-Marin, L. M., Breakspear, M., Martin, N. G., & Lupton, M. K. (2023). The Effect of Genetic Predisposition to Alzheimer’s Disease and Related Traits on Recruitment Bias in a Study of Cognitive Aging. Twin Research and Human Genetics, 26(3), 209–214. 10.1017/thg.2023.26

17. Hou, K., & Xu, X. (2023). Ambient temperatures associated with reduced cognitive function in older adults in China. Scientific Reports, 13(1), 17414. 10.1038/s41598-023-44776-2

18. IPCC. (2023). Climate Change 2021 – The Physical Science Basis: Working Group I Contribution to the Sixth Assessment Report of the Intergovernmental Panel on Climate Change (1st ed.). Cambridge University Press. 10.1017/9781009157896

19. Jeffrey, S. J., Carter, J. O., Moodie, K. B., & Beswick, A. R. (2001). Using spatial interpolation to construct a comprehensive archive of Australian climate data. Environmental Modelling & Software, 16(4), 309–330. 10.1016/S1364-8152(01)00008-1

20. Khan, A. M., Finlay, J. M., Clarke, P., Sol, K., Melendez, R., Judd, S., & Gronlund, C. J. (2021). Association between temperature exposure and cognition: A cross-sectional analysis of 20,687 aging adults in the United States. BMC Public Health, 21(1), 1484. 10.1186/s12889-021-11533-x

21. Lo, Y.-T. C., Su, W.-P., Mei, S.-H., Jou, Y.-Y., & Huang, H.-B. (2021). Association between ambient temperature and cognitive function in a community-dwelling elderly population: A repeated measurement study. BMJ Open, 11(12), e049160. 10.1136/bmjopen-2021-049160

22. Lupton, M. K., Robinson, G. A., Adam, R. J., Rose, S., Byrne, G. J., Salvado, O., Pachana, N. A., Almeida, O. P., McAloney, K., Gordon, S. D., Raniga, P., Fazlollahi, A., Xia, Y., Ceslis, A., Sonkusare, S., Zhang, Q., Kholghi, M., Karunanithi, M., Mosley, P. E., … Breakspear, M. (2021). A prospective cohort study of prodromal Alzheimer’s disease: Prospective Imaging Study of Ageing: Genes, Brain and Behaviour (PISA). NeuroImage: Clinical, 29, 102527. 10.1016/j.nicl.2020.102527

23. MacKinnon, J. G., & White, H. (1985). Some heteroskedasticity-consistent covariance matrix estimators with improved finite sample properties. Journal of Econometrics, 29(3), 305–325. 10.1016/0304-4076(85)90158-7

24. McMillan, J., Beavis, A., & Jones, F. L. (2009). The AUSEI06: A new socioeconomic index for Australia. Journal of Sociology, 45(2), 123–149. 10.1177/1440783309103342

25. Meade, R. D., Akerman, A. P., Notley, S. R., McGinn, R., Poirier, P., Gosselin, P., & Kenny, G. P. (2020). Physiological factors characterizing heat-vulnerable older adults: A narrative review. Environment International, 144, 105909. 10.1016/j.envint.2020.105909

26. Millyard, A., Layden, J. D., Pyne, D. B., Edwards, A. M., & Bloxham, S. R. (2020). Impairments to Thermoregulation in the Elderly During Heat Exposure Events. Gerontology and Geriatric Medicine, 6, 2333721420932432. 10.1177/2333721420932432

27. Minor, K., Bjerre-Nielsen, A., Jonasdottir, S. S., Lehmann, S., & Obradovich, N. (2022). Rising temperatures erode human sleep globally. One Earth, 5(5), 534–549. 10.1016/j.oneear.2022.04.008

28. Nairn, J., & Fawcett, R. (2014). The Excess Heat Factor: A Metric for Heatwave Intensity and Its Use in Classifying Heatwave Severity. International Journal of Environmental Research and Public Health, 12(1), 227–253. 10.3390/ijerph120100227

29. Nationwide House Energy Rating Scheme. (n.d.). NatHERS Climate Zone Postcodes. Retrieved June 14, 2025, from https://www.nathers.gov.au/climate-zone-postcodes?utm_source=chatgpt.com

30. Obradovich, N., Migliorini, R., Mednick, S. C., & Fowler, J. H. (2017). Nighttime temperature and human sleep loss in a changing climate. Science Advances, 3(5), e1601555. 10.1126/sciadv.1601555

31. Raymond, C., Matthews, T., & Horton, R. M. (2020). The emergence of heat and humidity too severe for human tolerance. Science Advances, 6(19), eaaw1838. 10.1126/sciadv.aaw1838

32. Sherwood, S. C., & Huber, M. (2010). An adaptability limit to climate change due to heat stress. Proceedings of the National Academy of Sciences, 107(21), 9552–9555. 10.1073/pnas.0913352107

33. Stroop, J. R. (1935). Studies of interference in serial verbal reactions. Journal of Experimental Psychology, 18(6), 643–662. 10.1037/h0054651

34. Stull, R. (2011). Wet-Bulb Temperature from Relative Humidity and Air Temperature. Journal of Applied Meteorology and Climatology, 50(11), 2267–2269. 10.1175/JAMC-D-11-0143.1

35. Thienel, R., Borne, L., Faucher, C., Behler, A., Robinson, G. A., Fripp, J., Giorgio, J., Ceslis, A., McAloney, K., Adsett, J., Galligan, D., Martin, N. G., Breakspear, M., & Lupton, M. K. (2024). Can an online battery match in-person cognitive testing in providing information about age-related cortical morphology? Brain Imaging and Behavior, 18(5), 1215–1225. 10.1007/s11682-024-00918-2

36. Wechsler, D. (1981). Manual for the Wechsler Adult Intelligence Scale—Revised. Psychological Corporation.

37. Wee, J., Tan, X. R., Gunther, S. H., Ihsan, M., Leow, M. K. S., Tan, D. S.-Y., Eriksson, J. G., & Lee, J. K. W. (2023). Effects of Medications on Heat Loss Capacity in Chronic Disease Patients: Health Implications Amidst Global Warming. Pharmacological Reviews, 75(6), 1140–1166. 10.1124/pharmrev.122.000782

38. Wijayanto, T., Toramoto, S., Maeda, Y., Son, S.-Y., Umezaki, S., & Tochihara, Y. (2017). Cognitive performance during passive heat exposure in Japanese males and tropical Asian males from Southeast Asian living in Japan. Journal of Physiological Anthropology, 36(1), 8. 10.1186/s40101-016-0124-4

39. Ye, Z., Li, X., Lang, H., Xin, J., Xu, H., & Fang, Y. (2024). Long-term effect of extreme temperature on cognitive function of middle-aged and older adults in China. International Journal of Geriatric Psychiatry, 39(2), e6063. 10.1002/gps.6063

40. Yeganeh, A. J., Reichard, G., McCoy, A. P., Bulbul, T., & Jazizadeh, F. (2018). Correlation of ambient air temperature and cognitive performance: A systematic review and meta-analysis. Building and Environment, 143, 701–716. 10.1016/j.buildenv.2018.07.002

41. Yin, B., Fang, W., Liu, L., Guo, Y., Ma, X., & Di, Q. (2024). Effect of extreme high temperature on cognitive function at different time scales: A national difference-in-differences analysis. Ecotoxicology and Environmental Safety, 275, 116238. 10.1016/j.ecoenv.2024.116238

42. Zander, K. K., Shalley, F., Taylor, A., Tan, G., & Dyrting, S. (2021). “Run air-conditioning all day”: Adaptation pathways to increasing heat in the Northern Territory of Australia. Sustainable Cities and Society, 74, 103194. 10.1016/j.scs.2021.103194

43. Zhao, Q., Wigmann, C., Areal, A. T., Altug, H., & Schikowski, T. (2021). Effect of non-optimum ambient temperature on cognitive function of elderly women in Germany. Environmental Pollution, 285, 117474. 10.1016/j.envpol.2021.117474

44. Zhou, W., Wang, Q., Li, R., Zhang, Z., Wang, W., Zhou, F., & Ling, L. (2023). The effects of heatwave on cognitive impairment among older adults: Exploring the combined effects of air pollution and green space. Science of The Total Environment, 904, 166534. 10.1016/j.scitotenv.2023.166534

