## Supplemental Material for "Warmer night-time temperatures are linked to poorer associative memory performance"

### Supplementary Material

#### *Temperature effects on extracted cognitive factors*

As an additional sensitivity analysis, we conducted an exploratory factor analysis of the cognitive task scores to assess whether temperature–cognition associations were evident at the level of broader data-driven cognitive factors. Participants with complete data across all included cognitive task scores were entered into the factor analysis, resulting in factor scores for 1,110 participants. The data were suitable for factor analysis. Bartlett's test of sphericity was significant, $\chi^2 = 1554.99$, $p < .001$, indicating that the cognitive-task correlation matrix was not an identity matrix and contained sufficient shared variance for factor analysis. Sampling adequacy was good, with an overall Kaiser–Meyer–Olkin measure of 0.854.

The number of factors to retain was guided by the eigenvalue structure of the cognitive-task correlation matrix. Three factors exceeded the Kaiser criterion of eigenvalue > 1 (Factor 1 = 3.128, Factor 2 = 1.105, Factor 3 = 1.025), while the fourth factor did not (eigenvalue = 0.935). We therefore retained three factors, interpreted as Memory, Visuospatial/Reasoning, and Verbal/Executive.

The retained three-factor solution explained 41.6% of the variance in the standardised cognitive task scores. After rotation, Factor 1 accounted for 17.7% of variance, Factor 2 for 11.0%, and Factor 3 for 12.9%. The mean communality across tasks was 0.416, indicating that the three-factor solution captured a moderate proportion of the shared variance across the cognitive battery. The factor loadings, communalities, and variance explained for the retained three-factor solution are reported in Supplementary Table S1.

The factor scores were then entered as outcomes in the same temperature-regression framework used for the primary task-level analyses. Across the three latent cognitive factors, there was no evidence that temperature exposure was associated with reduced-domain cognitive performance. Models tested $T_{max}$, $T_{min}$, $T_{max,percentile}$, $T_{min,percentile}$, $T_{w,max}$, and $T_{w,min}$. Season was included for absolute and wet-bulb temperature models, but not for percentile-based temperature models.

For Factor 1 ("Memory"), all temperature associations were negative, but none reached statistical significance. The strongest associations were observed for $T_{w,max}$ ($b = –0.0133$, 95% CI [–0.0290, 0.0024], $p = .096$) and $T_{w,min}$ ($b = –0.0108$, 95% CI [–0.0237, 0.0022], $p = .103$). For Factor 2 ("Visuospatial-Reasoning"), temperature coefficients were generally positive but non-significant. The strongest association was observed for $T_{w,max}$ ($b = 0.0122$, 95% CI [–0.0044, 0.0287], $p = .149$), with no evidence of association for percentile-based exposures or dry-bulb temperature measures. For Factor 3 ("Verbal-Executive"), no temperature variable was significantly associated with factor scores. Coefficients were close to zero for percentile-based exposures and small for absolute and wet-bulb temperature measures.

Overall, the exploratory factor analysis suggested that the cognitive battery could be summarised into three interpretable dimensions, but temperature exposure was not robustly associated with any of these reduced cognitive factors. This pattern supports the interpretation that the primary Paired Associates finding was not driven by a general reduction in cognitive performance or by a broad domain-level effect across the cognitive battery. Rather, the association appears task-specific, although this interpretation should remain cautious because Paired Associates was the only task primarily targeting episodic memory.

**Supplementary Table S1. Exploratory factor analysis of cognitive task scores.** Factor loadings from the three-factor exploratory factor analysis of standardised cognitive task scores are shown after oblimin rotation. The dominant loading for each task was used to guide factor interpretation. Factor 1 was interpreted as Memory, Factor 2 as Visuospatial/Reasoning, and Factor 3 as Verbal/Executive. Communalities indicate the proportion of variance in each task explained by the retained three-factor solution.

| **Cognitive task** | **Factor 1** | **Factor 2** | **Factor 3** | **Communality** |
|---|---|---|---|---|
| Monkey Ladder | **0.686** | 0.208 | –0.135 | 0.532 |
| Paired Associates | **0.659** | –0.257 | 0.058 | 0.504 |
| Spatial Span | **0.652** | 0.030 | 0.035 | 0.427 |
| Token Search | **0.521** | –0.040 | 0.152 | 0.296 |
| Spatial Planning | **0.460** | 0.262 | 0.084 | 0.287 |
| Rotations | 0.160 | **0.591** | 0.108 | 0.387 |
| Polygons | 0.106 | **0.516** | 0.221 | 0.326 |
| Feature Match | 0.400 | **0.412** | 0.039 | 0.331 |
| Odd One Out | 0.331 | **–0.453** | 0.175 | 0.345 |
| Digit Span | 0.053 | –0.243 | **0.729** | 0.594 |
| Grammatical Reasoning | 0.008 | 0.246 | **0.669** | 0.508 |
| Double Trouble | –0.053 | 0.180 | **0.649** | 0.456 |

**Supplementary Table S2. Sample size by cognitive test.**

| Test | N |
|---|---|
| Digit Span | 1613 |
| Double Trouble | 1584 |
| Feature Match | 1553 |
| Grammatical Reasoning | 1668 |
| Monkey Ladder | 1528 |
| Odd One Out | 1768 |
| Paired Associates | 1492 |
| Polygons | 1525 |
| Rotations | 1509 |
| Spatial Planning | 1563 |
| Spatial Span | 1545 |
| Token Search | 1604 |

**Supplementary Table S3. Participants per season and climate zone.**

| Climate zone | | Example locations | Spring | Summer | Autumn | Winter |
|---|---|---|---|---|---|---|
| 1 | Hot humid summer, warm winter | Darwin, Cairns, northern Australia | 11 | 5 | 5 | 3 |
| 2 | Warm humid summer, mild winter | Brisbane, coastal Queensland/ northern New South Wales | 233 | 150 | 158 | 131 |
| 3 | Hot dry summer, warm winter | Alice Springs, inland north | 4 | 6 | 4 | 7 |
| 4 | Hot dry summer, cool winter | inland New South Wales/ Victoria | 12 | 10 | 5 | 15 |
| 5 | Warm temperate | Sydney, Newcastle, Perth, Adelaide | 49 | 45 | 32 | 43 |
| 6 | Mild temperate | Melbourne, Ballarat, Albany | 155 | 136 | 104 | 103 |
| 7 | Cool temperate | Tasmania, south-eastern Victoria | 42 | 36 | 34 | 48 |
| 8 | Alpine | Alpine regions, Snowy Mountains | 0 | 0 | 0 | 0 |

**Supplementary Table S4. Regression Coefficients (b) for temperature predictors in quadratic model.**

| | Linear *b* | Quadratic $b^2$ | ΔAIC lin. – quad. | Adj. $R^2$ lin. | Adj. $R^2$ quad. |
|---|---|---|---|---|---|
| $\boldsymbol{T}_{\mathbf{max}}$ | | | | | |
| Digit Span | −0.010, p = .035 | −0.000, p = .442 | −1.38 | 0.045 | 0.044 |
| Double Trouble | −0.006, p = .152 | −0.000, p = .697 | −1.84 | 0.081 | 0.081 |
| Feature Match | −0.009, p = .054 | −0.000, p = .868 | −1.97 | 0.105 | 0.105 |
| Grammatical Reasoning | −0.000, p = .921 | −0.000, p = .865 | −1.97 | 0.125 | 0.125 |
| Monkey Ladder | 0.001, p = .753 | −0.000, p = .904 | −1.98 | 0.066 | 0.065 |
| Odd One Out | −0.001, p = .894 | 0.001, p = .109 | 0.70 | 0.001 | 0.002 |
| Paired Associates | −0.006, p = .169 | −0.000, p = .491 | −1.54 | 0.048 | 0.048 |
| Polygons | −0.004, p = .462 | 0.000, p = .789 | −1.92 | 0.055 | 0.055 |
| Rotations | −0.001, p = .896 | −0.000, p = .980 | −2.00 | 0.071 | 0.070 |
| Spatial Planning | −0.000, p = .945 | −0.000, p = .507 | −1.54 | 0.088 | 0.088 |
| Spatial Span | 0.000, p = .961 | −0.001, p = .088 | 0.64 | 0.069 | 0.070 |
| Token Search | −0.013, p = .004 | 0.000, p = .948 | −2.00 | 0.054 | 0.053 |
| $\boldsymbol{T}_{\mathbf{min}}$ | | | | | |
| Digit Span | −0.006, p = .262 | −0.001, p = .139 | 0.32 | 0.043 | 0.043 |
| Double Trouble | −0.006, p = .280 | −0.001, p = .217 | −0.45 | 0.081 | 0.081 |
| Feature Match | −0.003, p = .527 | 0.000, p = .519 | −1.53 | 0.103 | 0.103 |
| Grammatical Reasoning | 0.002, p = .667 | 0.000, p = .928 | −1.99 | 0.125 | 0.125 |
| Monkey Ladder | −0.001, p = .876 | −0.000, p = .891 | −1.98 | 0.066 | 0.065 |
| Odd One Out | 0.000, p = .959 | 0.001, p = .201 | −0.37 | 0.001 | 0.001 |
| Paired Associates | −0.017, p = .001 | −0.001, p = .357 | −1.19 | 0.053 | 0.053 |
| Polygons | −0.001, p = .874 | −0.000, p = .964 | −2.00 | 0.055 | 0.054 |
| Rotations | −0.002, p = .678 | 0.000, p = .989 | −2.00 | 0.071 | 0.070 |
| Spatial Planning | −0.006, p = .239 | −0.000, p = .790 | −1.94 | 0.089 | 0.089 |
| Spatial Span | 0.004, p = .496 | −0.001, p = .132 | 0.14 | 0.070 | 0.070 |
| Token Search | −0.014, p = .010 | 0.000, p = .535 | −1.67 | 0.053 | 0.052 |
| $\boldsymbol{T}_{\mathbf{w,max}}$ | | | | | |
| Digit Span | −0.013, p = .064 | −0.001, p = .382 | −1.13 | 0.044 | 0.044 |
| Double Trouble | −0.011, p = .101 | −0.001, p = .242 | −0.67 | 0.082 | 0.082 |
| Feature Match | −0.008, p = .269 | 0.000, p = .969 | −2.00 | 0.104 | 0.103 |
| Grammatical Reasoning | −0.000, p = .985 | 0.000, p = .854 | −1.97 | 0.125 | 0.125 |
| Monkey Ladder | −0.001, p = .925 | −0.000, p = .810 | −1.94 | 0.066 | 0.065 |
| Odd One Out | −0.001, p = .903 | 0.001, p = .529 | −1.59 | 0.001 | 0.000 |
| Paired Associates | −0.017, p = .013 | −0.001, p = .213 | −0.51 | 0.051 | 0.051 |
| Polygons | −0.002, p = .767 | 0.001, p = .462 | −1.40 | 0.055 | 0.055 |
| Rotations | −0.001, p = .881 | −0.001, p = .569 | −1.64 | 0.071 | 0.070 |
| Spatial Planning | −0.005, p = .442 | −0.001, p = .400 | −1.30 | 0.089 | 0.088 |
| Spatial Span | 0.001, p = .900 | −0.001, p = .207 | −0.31 | 0.069 | 0.070 |

| | | | | | |
|---|---|---|---|---|---|
| Token Search | −0.016, p = .023 | 0.000, p = .852 | −1.97 | 0.052 | 0.052 |
| ***T*****w,min** | | | | | |
| Digit Span | −0.007, p = .257 | −0.001, p = .079 | 1.31 | 0.043 | 0.044 |
| Double Trouble | −0.007, p = .209 | −0.001, p = .147 | 0.06 | 0.081 | 0.081 |
| Feature Match | −0.002, p = .758 | 0.000, p = .827 | −1.94 | 0.103 | 0.102 |
| Grammatical Reasoning | 0.002, p = .767 | −0.000, p = .697 | −1.85 | 0.125 | 0.125 |
| Monkey Ladder | −0.002, p = .715 | −0.000, p = .621 | −1.73 | 0.066 | 0.065 |
| Odd One Out | 0.000, p = .977 | 0.001, p = .411 | −1.29 | 0.001 | 0.000 |
| Paired Associates | −0.018, p = .001 | −0.001, p = .278 | −0.85 | 0.053 | 0.053 |
| Polygons | 0.001, p = .897 | 0.000, p = .878 | −1.98 | 0.055 | 0.054 |
| Rotations | −0.002, p = .725 | −0.000, p = .512 | −1.56 | 0.071 | 0.070 |
| Spatial Planning | −0.008, p = .172 | −0.001, p = .454 | −1.47 | 0.089 | 0.089 |
| Spatial Span | 0.003, p = .586 | −0.001, p = .147 | 0.05 | 0.069 | 0.070 |
| Token Search | −0.011, p = .060 | 0.000, p = .739 | −1.90 | 0.051 | 0.051 |
| ***T*****max,percentile** | | | | | |
| Digit Span | −0.000, p = .640 | −0.000, p = .198 | −0.20 | 0.041 | 0.041 |
| Double Trouble | 0.001, p = .404 | −0.000, p = .186 | −0.24 | 0.070 | 0.071 |
| Feature Match | −0.001, p = .169 | −0.000, p = .366 | −1.13 | 0.104 | 0.104 |
| Grammatical Reasoning | −0.001, p = .317 | −0.000, p = .466 | −1.47 | 0.128 | 0.128 |
| Monkey Ladder | 0.000, p = .859 | 0.000, p = .612 | −1.73 | 0.066 | 0.066 |
| Odd One Out | −0.001, p = .138 | 0.000, p = .977 | −2.00 | 0.001 | 0.000 |
| Paired Associates | 0.000, p = .568 | −0.000, p = .733 | −1.88 | 0.044 | 0.043 |
| Polygons | −0.001, p = .414 | 0.000, p = .084 | 1.18 | 0.056 | 0.058 |
| Rotations | 0.000, p = .551 | −0.000, p = .624 | −1.76 | 0.072 | 0.072 |
| Spatial Planning | −0.000, p = .947 | 0.000, p = .973 | −2.00 | 0.089 | 0.088 |
| Spatial Span | −0.000, p = .776 | 0.000, p = .668 | −1.81 | 0.070 | 0.069 |
| Token Search | −0.000, p = .689 | −0.000, p = .111 | 0.68 | 0.048 | 0.049 |
| ***T*****min,percentile** | | | | | |
| Digit Span | −0.000, p = .903 | −0.000, p = .316 | −0.94 | 0.041 | 0.041 |
| Double Trouble | −0.000, p = .790 | −0.000, p = .458 | −1.46 | 0.069 | 0.069 |
| Feature Match | −0.000, p = .631 | 0.000, p = .368 | −1.20 | 0.104 | 0.104 |
| Grammatical Reasoning | −0.000, p = .883 | −0.000, p = .951 | −2.00 | 0.126 | 0.126 |
| Monkey Ladder | −0.000, p = .857 | 0.000, p = .376 | −1.22 | 0.066 | 0.066 |
| Odd One Out | −0.000, p = .601 | 0.000, p = .029 | 2.57 | 0.000 | 0.002 |
| Paired Associates | −0.002, p = .002 | −0.000, p = .586 | −1.71 | 0.050 | 0.049 |
| Polygons | −0.000, p = .928 | 0.000, p = .777 | −1.92 | 0.055 | 0.055 |
| Rotations | 0.000, p = .648 | −0.000, p = .068 | 1.49 | 0.072 | 0.074 |
| Spatial Planning | 0.000, p = .991 | −0.000, p = .468 | −1.46 | 0.088 | 0.088 |
| Spatial Span | 0.001, p = .391 | 0.000, p = .298 | −0.94 | 0.070 | 0.070 |
| Token Search | −0.000, p = .908 | −0.000, p = .403 | −1.27 | 0.048 | 0.047 |

**Supplementary Table S5. Linear model regression coefficients (*b*) for temperature predictors across cognitive tests.** Each coefficient represents the change in z-scored cognitive performance per unit increase in the temperature variable, from separate models adjusting for age, sex, education, SEI, and climate zone. p-values were adjusted for False Discovery Rate (FDR) globally.

| | *b* | SE | CI | *p* | $p^{FDR}$ |
|---|---|---|---|---|---|
| $T_{max}$ | | | | | |
| Digit Span | -0.0102 | 0.0048 | [-0.0196, -0.0007] | 0.04 | 0.32 |
| Double Trouble | -0.0065 | 0.0045 | [-0.0154, 0.0024] | 0.15 | 0.73 |
| Feature Match | -0.0089 | 0.0046 | [-0.0179, 0.0001] | 0.05 | 0.42 |
| Grammatical Reasoning | -0.0004 | 0.0042 | [-0.0086, 0.0078] | 0.92 | 0.99 |
| Monkey Ladder | 0.0015 | 0.0047 | [-0.0077, 0.0107] | 0.75 | 0.99 |
| Odd One Out | -0.0006 | 0.0043 | [-0.0091, 0.0079] | 0.89 | 0.99 |
| Paired Associates | -0.0062 | 0.0045 | [-0.0151, 0.0027] | 0.17 | 0.73 |
| Polygons | -0.0036 | 0.0049 | [-0.0133, 0.0060] | 0.46 | 0.99 |
| Rotations | -0.0006 | 0.0048 | [-0.0100, 0.0087] | 0.90 | 0.99 |
| Spatial Planning | -0.0003 | 0.0044 | [-0.0090, 0.0084] | 0.94 | 0.99 |
| Spatial Span | 0.0002 | 0.0046 | [-0.0088, 0.0093] | 0.96 | 0.99 |
| Token Search | -0.0132 | 0.0046 | [-0.0222, -0.0047] | 0.004 | 0.07 |
| $T_{min}$ | | | | | |
| Digit Span | -0.0063 | 0.0056 | [-0.0174, 0.0047] | 0.26 | 0.88 |
| Double Trouble | -0.00569 | 0.00527 | [-0.01602, 0.00464] | 0.28 | 0.88 |
| Feature Match | -0.00340 | 0.00537 | [-0.01394, 0.00714] | 0.53 | 0.99 |
| Grammatical Reasoning | 0.00214 | 0.00498 | [-0.00762, 0.011902] | 0.67 | 0.99 |
| Monkey Ladder | -0.00091 | 0.00583 | [-0.01235, 0.01053] | 0.88 | 0.99 |
| Odd One Out | 0.00026 | 0.00519 | [-0.00991, 0.01044] | 0.96 | 0.99 |
| Paired Associates | -0.01682 | 0.00522 | [-0.02705, -0.00659] | 0.001 | **0.046** |
| Polygons | -0.00088 | 0.00557 | [-0.01182, 0.01005] | 0.87 | 0.99 |
| Rotations | -0.00234 | 0.00564 | [-0.01340, 0.00872] | 0.68 | 0.99 |
| Spatial Planning | -0.00627 | 0.00532 | [-0.01672, 0.00417] | 0.24 | 0.88 |
| Spatial Span | 0.00362 | 0.00532 | [-0.00681, 0.01405] | 0.50 | 0.99 |
| Token Search | -0.01357 | 0.00529 | [-0.02400, -0.00319] | 0.010 | 0.15 |
| $T_{w,max}$ | | | | | |
| Digit Span | -0.01327 | 0.00716 | [-0.02731, 0.00077] | 0.06 | 0.42 |

| | | | | | |
|---|---|---|---|---|---|
| Double Trouble | -0.01104 | 0.00672 | [-0.02423, 0.00214] | 0.10 | 0.60 |
| Feature Match | -0.00753 | 0.00681 | [-0.02088, 0.00583] | 0.27 | 0.88 |
| Grammatical Reasoning | -0.00012 | 0.00626 | [-0.01240, 0.01216] | 0.98 | 0.99 |
| Monkey Ladder | -0.00067 | 0.00713 | [-0.01465, 0.01332] | 0.93 | 0.99 |
| Odd One Out | -0.00078 | 0.00644 | [-0.01341, 0.01184] | 0.90 | 0.99 |
| Paired Associates | -0.01656 | 0.00665 | [-0.02961, -0.00351] | 0.013 | 0.15 |
| Polygons | -0.00211 | 0.00714 | [-0.01611, 0.01189] | 0.76 | 0.99 |
| Rotations | -0.00107 | 0.00712 | [-0.01503, 0.01290] | 0.88 | 0.99 |
| Spatial Planning | -0.00517 | 0.00672 | [-0.01836, 0.00801] | 0.44 | 0.99 |
| Spatial Span | 0.00087 | 0.00693 | [-0.01273, 0.01447] | 0.90 | 0.99 |
| Token Search | -0.01554 | 0.00681 | [-0.02890, -0.00218] | 0.023 | 0.23 |
| $T_{w,min}$ | | | | | |
| Digit Span | -0.00661 | 0.00583 | [-0.01805, 0.00483] | 0.26 | 0.88 |
| Double Trouble | -0.00689 | 0.00548 | [-0.01764, 0.00387] | 0.21 | 0.84 |
| Feature Match | -0.00173 | 0.00562 | [-0.01276, 0.00929] | 0.76 | 0.99 |
| Grammatical Reasoning | 0.00155 | 0.00522 | [-0.00869, 0.01179] | 0.77 | 0.99 |
| Monkey Ladder | -0.00223 | 0.00610 | [-0.01420, 0.00974] | 0.71 | 0.99 |
| Odd One Out | 0.00015 | 0.00530 | [-0.01024, 0.01054] | 0.98 | 0.99 |
| Paired Associates | -0.01799 | 0.00547 | [-0.02873, -0.00725] | 0.001 | **0.046** |
| Polygons | 0.00074 | 0.00572 | [-0.01048, 0.01197] | 0.90 | 0.99 |
| Rotations | -0.00203 | 0.00578 | [-0.01336, 0.00930] | 0.73 | 0.99 |
| Spatial Planning | -0.00765 | 0.00559 | [-0.01862, 0.00333] | 0.17 | 0.73 |
| Spatial Span | 0.00306 | 0.00562 | [-0.00795, 0.01408] | 0.59 | 0.99 |
| Token Search | -0.01051 | 0.00559 | [-0.02149, 0.00046] | 0.06 | 0.42 |
| $T_{max,percentile}$ | | | | | |
| Digit Span | -0.00038 | 0.00082 | [-0.00199, 0.00123] | 0.64 | 0.99 |
| Double Trouble | 0.00065 | 0.00078 | [-0.00087, 0.00217] | 0.40 | 0.99 |
| Feature Match | -0.00105 | 0.00077 | [-0.00256, 0.00045] | 0.17 | 0.73 |
| Grammatical Reasoning | -0.00075 | 0.00074 | [-0.00221, 0.00071] | 0.32 | 0.95 |
| Monkey Ladder | 0.00014 | 0.00080 | [-0.00142, 0.00171] | 0.86 | 0.99 |
| Odd One Out | -0.00113 | 0.00076 | [-0.00263, 0.00036] | 0.14 | 0.73 |
| Paired Associates | 0.00047 | 0.00082 | [-0.00114, 0.00208] | 0.57 | 0.99 |
| Polygons | -0.00068 | 0.00083 | [-0.00230, 0.00095] | 0.41 | 0.99 |
| Rotations | 0.00048 | 0.00080 | [-0.00109, 0.00205] | 0.55 | 0.99 |
| Spatial Planning | -0.00005 | 0.00077 | [-0.00157, 0.00147] | 0.95 | 0.99 |

| | | | | | |
|---|---|---|---|---|---|
| Spatial Span | -0.00023 | 0.00081 | [-0.00182, 0.00136] | 0.78 | 0.99 |
| Token Search | -0.00033 | 0.00081 | [-0.00192, 0.00127] | 0.69 | 0.99 |
| ***T*min,percentile** | | | | | |
| Digit Span | -0.00010 | 0.00081 | [-0.00169, 0.00149] | 0.90 | 0.99 |
| Double Trouble | -0.00020 | 0.00077 | [-0.00170, 0.00130] | 0.79 | 0.99 |
| Feature Match | -0.00037 | 0.00077 | [-0.00189, 0.00115] | 0.63 | 0.99 |
| Grammatical Reasoning | -0.00011 | 0.00073 | [-0.00153, 0.00132] | 0.88 | 0.99 |
| Monkey Ladder | -0.00014 | 0.00079 | [-0.00170, 0.00141] | 0.86 | 0.99 |
| Odd One Out | -0.00039 | 0.00074 | [-0.00185, 0.00107] | 0.60 | 0.99 |
| Paired Associates | -0.00246 | 0.00079 | [-0.00402, -0.00090] | 0.002 | **0.048** |
| Polygons | -0.00007 | 0.00078 | [-0.00161, 0.00147] | 0.93 | 0.99 |
| Rotations | 0.00036 | 0.00078 | [-0.00117, 0.00189] | 0.65 | 0.99 |
| Spatial Planning | 0.00001 | 0.00077 | [-0.00150, 0.00152] | 0.99 | 0.99 |
| Spatial Span | 0.00065 | 0.00076 | [-0.00083, 0.00213] | 0.39 | 0.99 |
| Token Search | -0.00009 | 0.00078 | [-0.00163, 0.00145] | 0.91 | 0.99 |

**Supplementary Figure S1. Distribution of environmental temperature exposures by season and climate zone.** Black diamonds and error bars show group means and standard deviation.

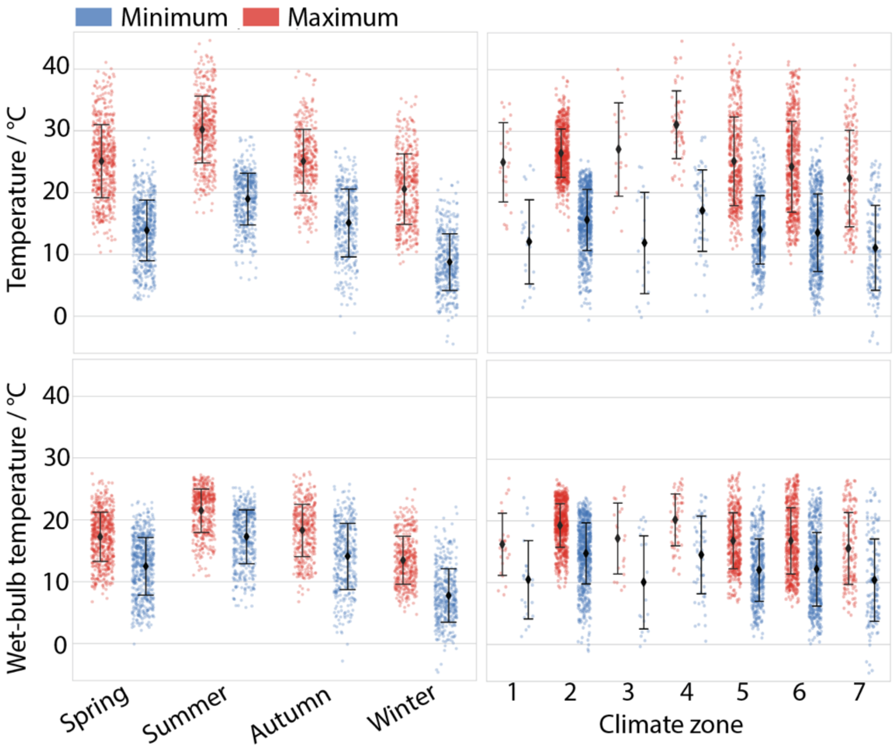
